# Computational modelling of COVID-19: A study of compliance and superspreaders

**DOI:** 10.1101/2021.05.12.21257079

**Authors:** Faith Lee, Maria Perez Ortiz, John Shawe-Taylor

## Abstract

**Background:** The success of social distancing implementations of severe acute respiratory syndrome coronavirus 2 (SARS-Cov-2) depends heavily on population compliance. Mathematical modelling has been used extensively to assess the rate of viral transmission from behavioural responses. Previous epidemics of SARS-Cov-2 have been characterised by superspreaders, a small number of individuals who transmit a disease to a large group of individuals, who contribute to the stochasticity (or randomness) of transmission compared to other pathogens such as Influenza. This growing evidence proves an urgent matter to understand transmission routes in order to target and combat outbreaks.

**Objective:** To investigate the role of superspreaders in the rate of viral transmission with various levels of compliance.

**Method:** A SEIRS inspired social network model is adapted and calibrated to observe the infected links of a general population with and without superspreaders on four compliance levels. Local and global connection parameters are adjusted to simulate close contact networks and travel restrictions respectively and each performance assessed. The mean and standard deviation of infections with superspreaders and non-superspreaders were calculated for each compliance level.

**Results:** Increased levels of compliance of superspreaders proves a significant reduction in infections. Assuming long-lasting immunity, superspreaders could potentially slow down the spread due to their high connectivity.

**Discussion:** The main advantage of applying the network model is to capture the heterogeneity and locality of social networks, including the role of superspreaders in epidemic dynamics. The main challenge is the immediate attention on social settings with targeted interventions to tackle superspreaders in future empirical work.

**Conclusion:** Superspreaders play a central role in slowing down infection spread following compliance guidelines. It is crucial to adjust social distancing measures to prevent future outbreaks accompanied by population-wide testing and effective tracing.

## 1. Introduction

Since the outbreak of SARS-Cov-2, different countries have implemented various social restriction measures in order to combat and reduce the spread of the virus [1]. Non-pharmaceutical interventions (NPIs) such as wearing masks, staying home, restrictions on public gatherings are advised to reduce the spread [2]. In order to prevent community outbreaks, contact tracing, isolation and quarantine are carried out for the containment of spread [3]. These strategies are strongly dependent on the compliance of individuals, in which compliance represents the strength of adherence to strict distancing guidelines.

Mathematical and statistical modelling are invaluable epidemiological tools. They allow public health officials to conduct virtual experiments to evaluate the efficacy of control strategies that might be practically infeasible [4]. Specifically, experiments evaluating the value of control strategies are impossible in practice, as it is unethical to introduce diseases intentionally into populations or withhold life-saving interventions for solely scientific research. They are thus of great importance, enabling a principled evaluation of strategies such as vaccination and quarantine; a way around the difficulty of studying such interventions in the real world.

In this report, a novel SEIRS Network model is adapted to evaluate simulated levels of compliance for both local and global social connectivity for superspreaders. A superspreader is an individual who transmits an infectious disease to an unexpectedly large group of individuals. In fact, previous work has shown a super-linear relationship between the number of contacts and duration, indicating the possibility that superspreaders need to be defined not only in the number of connections but also in intensity [5]. This means that with more distinctive interactions per individual, the higher the average duration of those interactions. Using the SEIRS Network model as an extension to the classic SEIR model provides an advantage of distinguishing infectious individuals (nodes) through social contacts and analysing behavioural responses. The proposed findings are that with higher levels of compliance, the rate of viral transmission reduces along with the number of infections in the population. The rate of spread also depends heavily on the compliance of superspreaders as they have a prominent effect on the infectivity. It is critical that drastic measures are taken to target social settings and limit the movement and spread of superspreaders.

## 2. Methods

### 2.1 SEIR model

The standard SEIR model assumes uniform mixing and homogeneity of infectious individuals with the wider population [6]. Each individual (or node) is associated with a current state: S (susceptible), E (exposed), I (infected) or R (recovered). The basic reproduction rate R_0_, which is the number of secondary infections infected in a susceptible population, is derived by β/γ. Current policies have failed to consider the heterogeneity and potential of superspreaders. By using the SEIRS Network model (discussed later), we analyse and evaluate scale-free network connections to suggest the undeniable role of superspreaders in affecting viral transmission. Scale-free networks (also named Barabasi-Albert graph) are social graphs marked by a highly skewed distribution of contacts in which most of the nodes are weakly connected, but only a small proportion of nodes having very high connectivity like superspreaders [7]. This source of heterogeneity is crucial in epidemiological simulations and allows policy interventions to be implemented more effectively to slow the spread of viral transmission.

**Explanation of parameters:**

**β**: rate of transmission

**σ**: rate of progression from exposed state to infectious state

**γ**: rate of recovery

N: Total population (S + E + I + R)

### 2.2 SEIRS Network model

We extend the compartmental SEIR model to stochastic networks^2^. A modified scale-free graph is simulated with 10000 nodes and an estimate of 12 connections per node on average to prototype parameters and scenarios as demonstrated by previous literature [8]. At a given time, an individual makes contact with a subset of random individuals from their set of close contacts (local interactions) with a certain probability and with a subset of individuals outside of their network (global interactions) with a different probability.

More specifically, in our model, we consider two different types of social interactions [9]:

a. Local interactions (p_local), i.e. with close contacts. These are individuals with whom one has repeated, sustained, and/or physical interactions on a regular basis. Examples include housemates, family members, close friends.
b. Global interactions (p_global), i.e. with casual contacts. These are individuals with whom one has incidental, or brief contact on an infrequent basis. Examples include one-off or random acquaintances at the grocery store, on transit or in an elevator. Global interactions are represented in the models in the form of a parallel mode of mean-field global transmission.

### 2.3 Experimental Conditions

Overall, we categorise individuals into 4 core levels of compliances. The parameter values are suggested to simulate how a population would interact given the restrictions:

- Full (or 100%) compliance: complying strictly with guidelines and minimising local and global connectivity
- Partial compliance is divided into two parts:
  - (i): complying with travel restrictions, meaning reducing global contacts whilst maintaining close family and friends contact.
  - (ii): complying with close contacts, meaning reducing local network but increasing rare acquaintances in global connectivity.

- No (or 0%) compliance: Ignoring social distancing guidelines and maximising local and global connectivity.

Table 1 reports the simulation parameters of local and global connectivity. We chose these values to represent the probability of social interactions on local and global transmission. Both partial compliances have the same probability of social interactions; p_local + p_global will yield the same probability.

**Table 1.**
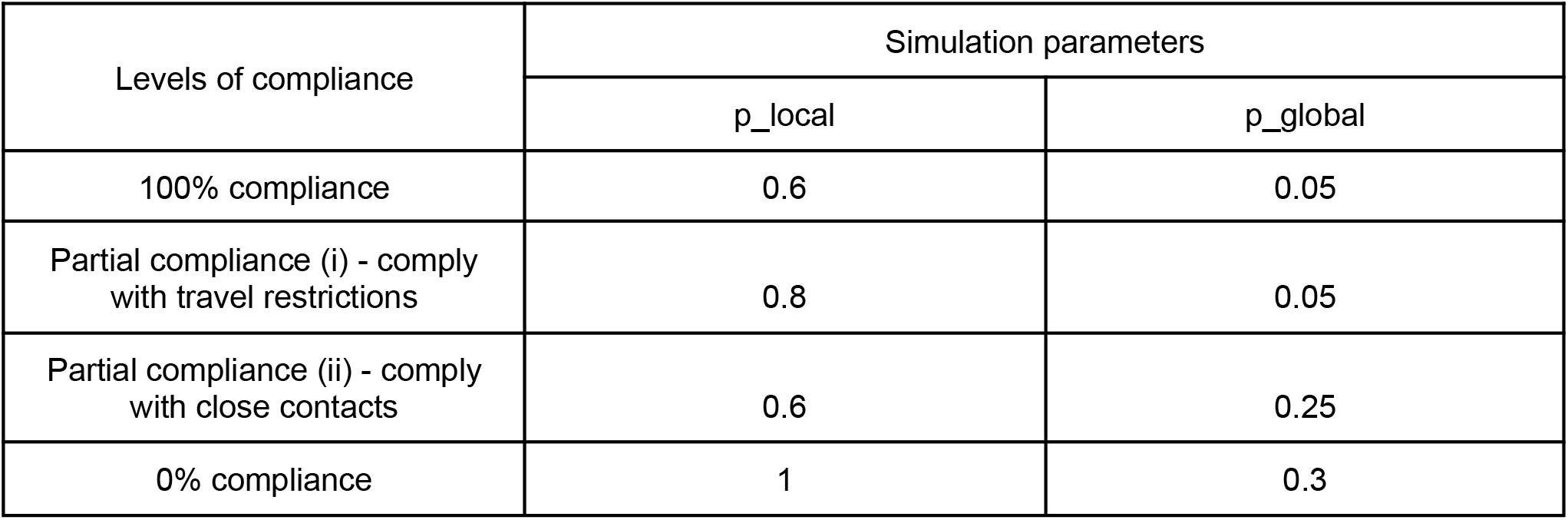
Probability of simulation parameters for p_local and p_global.

### 2.4 Degree distributions of homogeneous and heterogeneous populations

Histograms are plotted below to demonstrate the number of nodes (individuals) and the degree distribution of homogeneous (Fig.6A) and heterogeneous (Fig.6B) populations. Homogeneous represents a population without superspreaders, and each individual has an approximately uniform mixing of social contacts. Heterogeneous, however, includes superspreaders which have high connectivity. The degree distribution is defined as the number of edges (or connections) that each node has to other nodes. It can also be seen as the probability distribution of degrees throughout the contact network.

**Figure 1.**
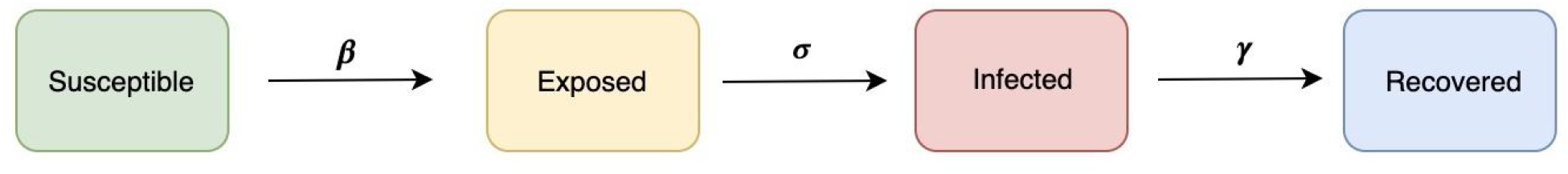
SEIR model with the progression of four states.

**Figure 2.**
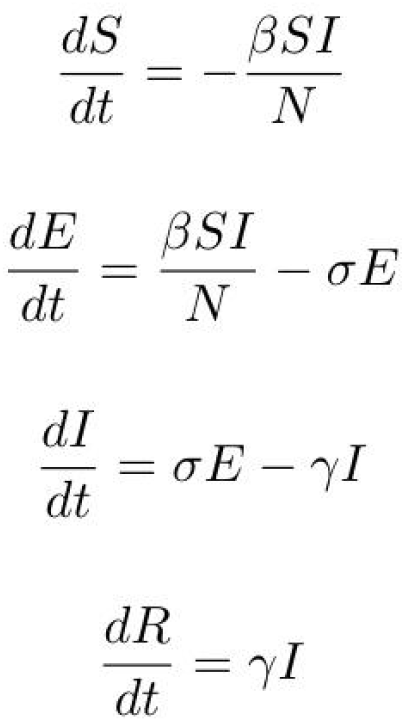
The viral transmission can be explained using a set of nonlinear differential equations.

**Figure 3.**
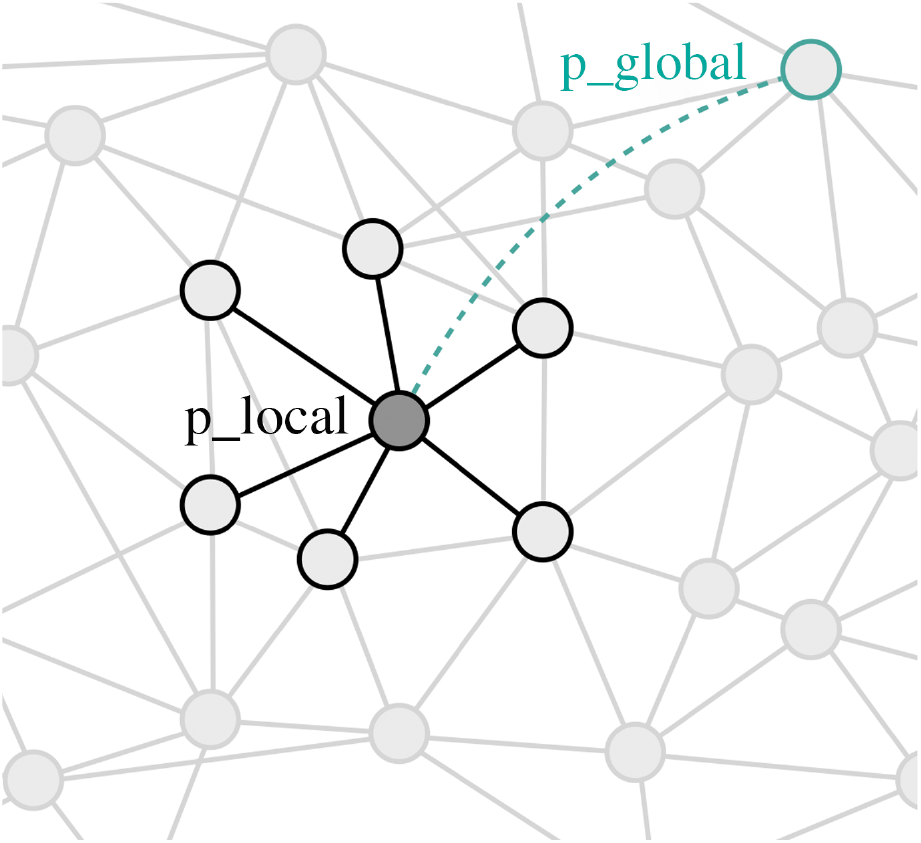
Contact network of parameters p_local and p_global used for simulations [9]. A dark grey circle (G) represents an individual or node, and their interactions (edges). The nodes situated adjacent to G represent the individual’s close contacts. The nodes further away represent global contacts of individuals.

**Figure 4.**
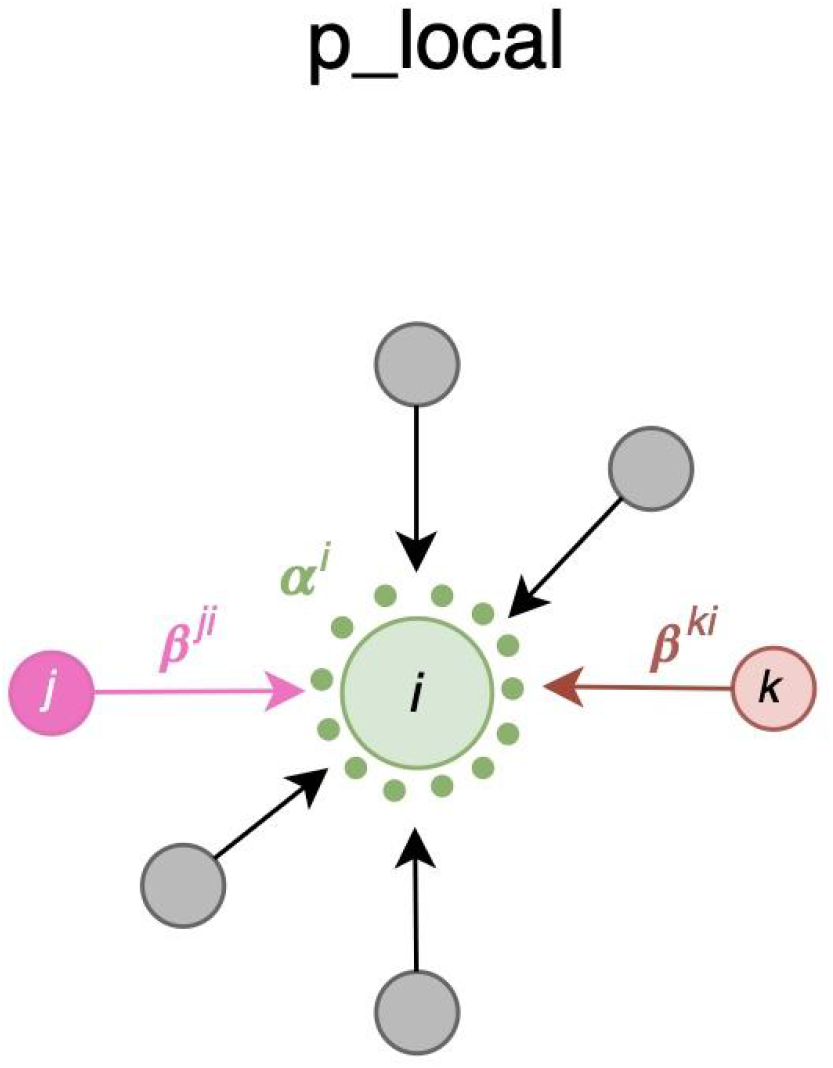
Diagram showing contact nodes and transmission for local connectivity [9]. Given a susceptible (α) individual (i), the transmissibility β is considered from infected node j to susceptible node i, providing the transmissibility weight βji. The same explanation goes to infected node k.

**Figure 5.**
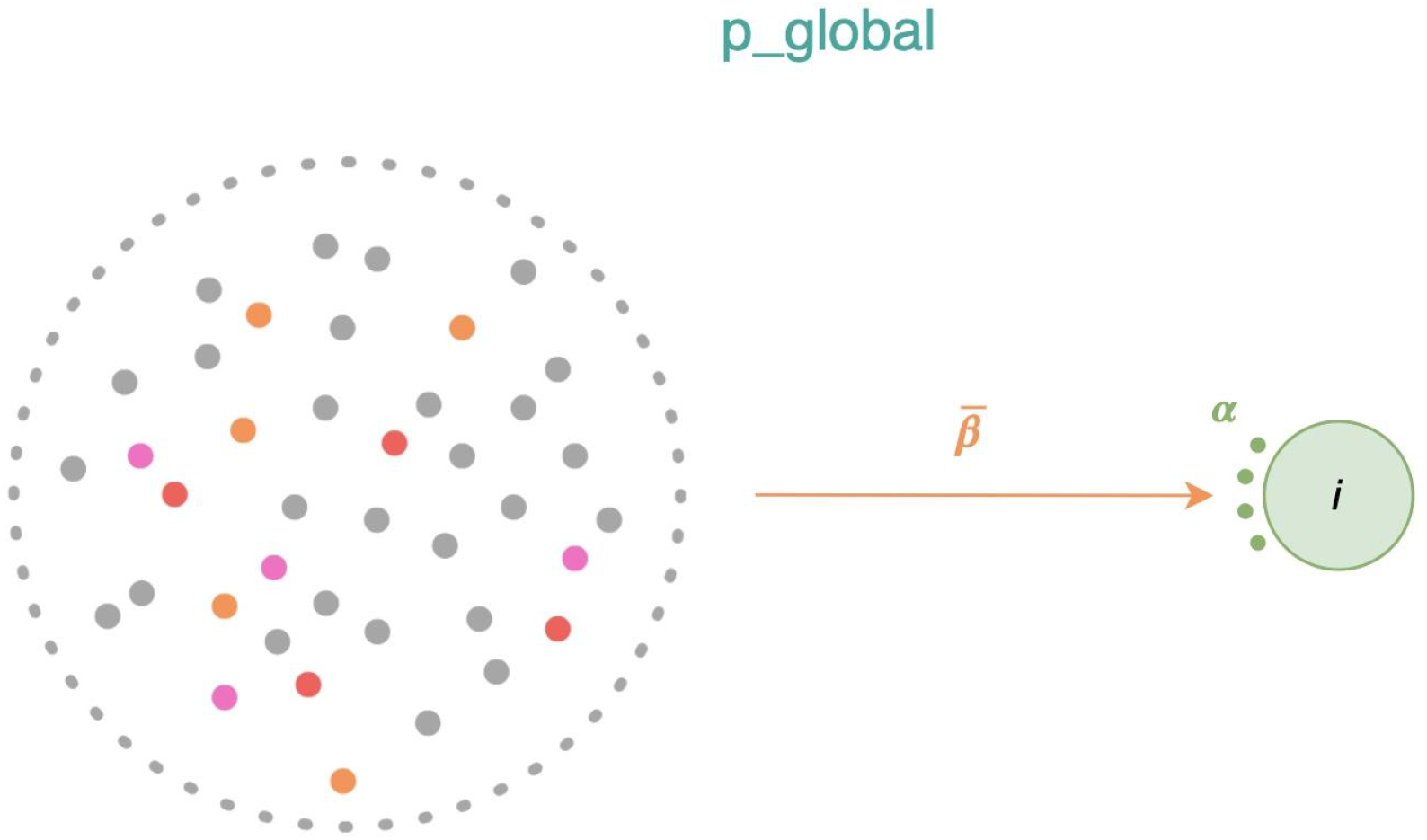
Diagram showing contact nodes and transmission for global network [9]. The global contacts are considered as a one-off encounter with unknown individuals in public. Each node is likely to interact with every node in the population, thus the probability for a susceptible individual (i) is provided by the mean transmissibility β.

**Figure 6.**
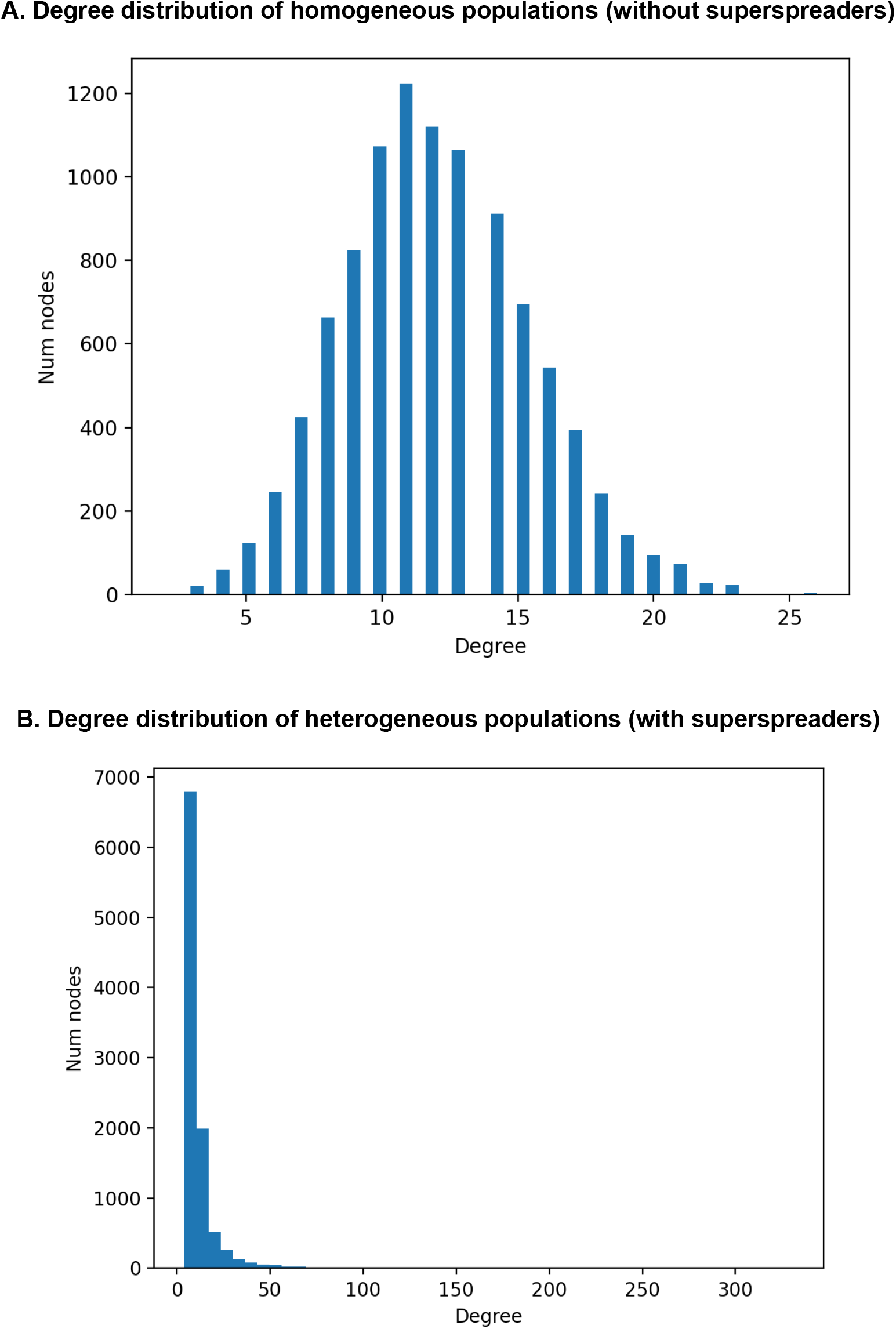
Comparison of degree distribution of homogeneous and heterogeneous populations. (A) Nodes and degree distribution of homogeneous populations. A normal distribution is observed. (B) Nodes and degree distribution of heterogeneous populations. A skewed distribution is observed.

Simulations with homogeneous populations are explained by the Erdos-Renyi network whilst scale-free networks describe heterogeneous populations [10]. Erdos-Renyi, or random homogeneous networks, are the underlying assumption of most epidemiological models as the degree distribution is approximated by Poisson and peaks around the average, denoting a statistical homogeneity of nodes. Whereas scale-free networks contain broad heterogeneity which results in a power-law distribution, indicating effects of superspreaders.

Figure 6A shows a seemingly general (or normal) distribution of the number of nodes in the interactions of non-superspreaders. The histogram peaks around 1200 individuals with a degree of 11. The maximum degree shown here is 25, which is much lower compared to figure 6B, which has a maximum degree of 300. This demonstrates the low connectivity of non-superspreaders.

Figure 6B shows a highly skewed distribution of the degree distribution in a scale-free heterogeneous population. The histogram peaks around 6500 individuals which is 5 times higher than that of figure 6A. The maximum degree that can be reached is around 300 which shows the high connectivity of superspreaders. This amount of connections effectively means that superspreaders get infected and immunised quickly.

## 3. Results

Simulations for both homogeneous and heterogeneous populations are run 30 times. We then report the mean and standard deviation (s.d) for the total percentage of infected individuals and percentage of individuals infected at peak in the tables below. It is important to note that all the simulations in this work entail exactly the same amount of social interactions per time step, independently of homogeneous or heterogeneous populations [11], with exception of the different levels of compliance, in which the values of parameters p_local and p_global set the amount and type of social interactions.

### 3.1 Compliance levels of homogeneous populations (without superspreaders)

The first simulation is conducted with homogeneous populations. The lowest mean total percentage infected and at peak is observed with full compliance and highest with no compliance as expected. Partial compliance (i) shows a lowered infected mean at peak and total infections than partial compliance (ii). This shows that not all social interactions are equal, in particular, global interactions (partial compliance (ii)) increase the spread significantly when compared to local interactions (partial compliance (i)). The same is applicable to the peak of infections. In addition, the s.d for the total percentage of infected is the highest with full compliance, followed by partial compliance (i), (ii) and no compliance, suggesting higher stochasticity. However, low s.d is seen in all results of percentage of peak infections indicating a low spread from the mean.

### 3.2 Compliance levels of heterogeneous populations (with superspreaders)

A second simulation is conducted with heterogeneous populations. It is anticipated that full compliance has the lowest infections and no compliance entails the highest infections. A particularly striking observation is that a heterogeneous population leads, in all cases except for 100% compliance, to a reduction of the spread but a higher peak of infections, when compared to a homogeneous population (i.e. compared to the results in Table 2). The hypothesis is that, assuming long-lasting sterile immunity, superspreaders usually get infected quickly and then become immune, and given that they are highly connected they help reduce the spread overall, but since the spread advances quicker the peak is higher [10], as shown in previous literature. The results also show that partial compliance (i) now shows similar total infections and infected nodes at peak than partial compliance (ii). It seems that in this case, superspreaders are so connected that the spread of the pathogen is similar independently of the type of social interactions (global or local, as indicated by the two different partial compliances).

**Table 2.**
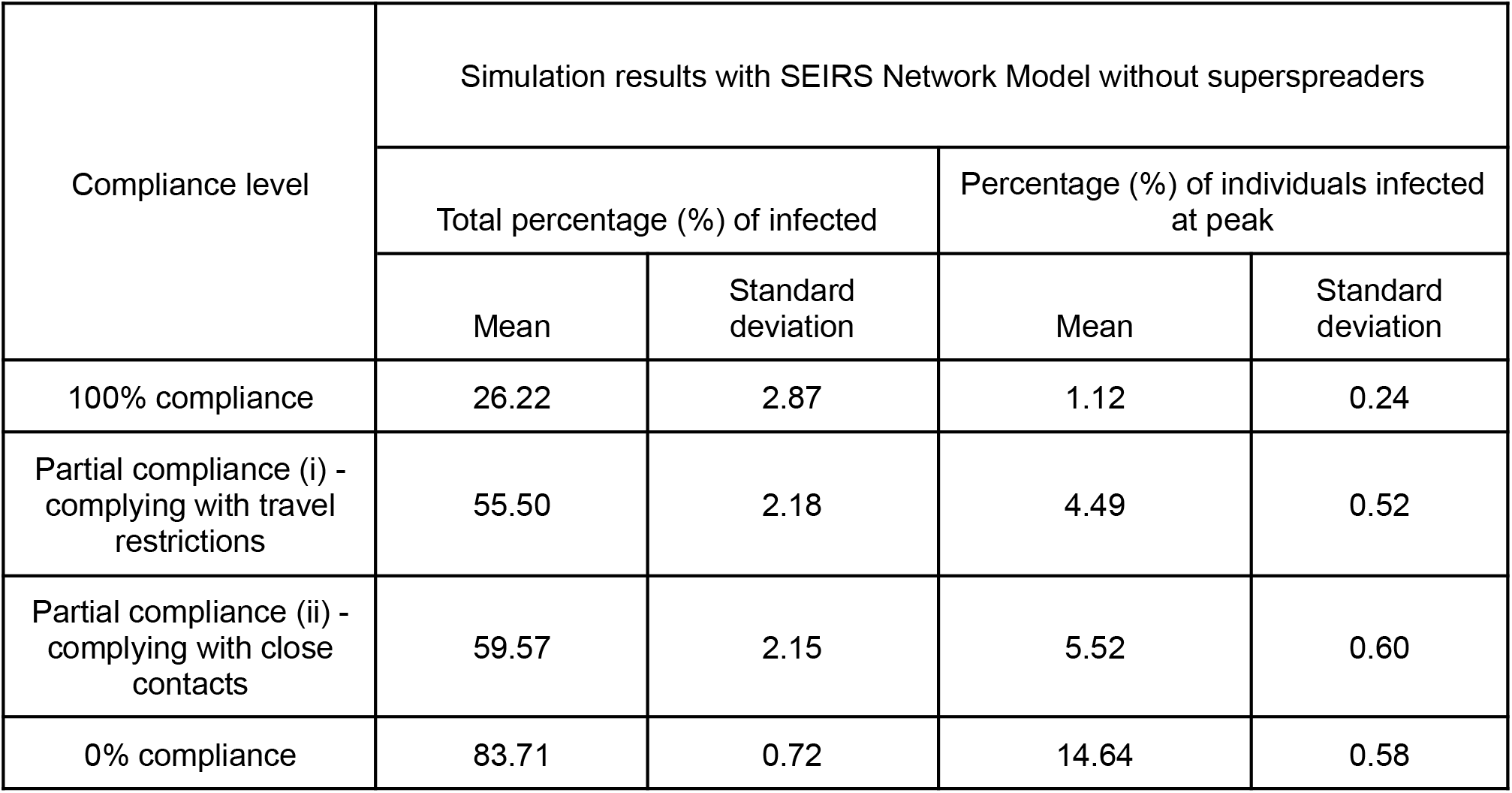
Mean and standard deviation of simulation results for total percentage of infected individuals and percentage of individuals infected at peak of homogeneous populations (without superspreaders).

**Table 3.**
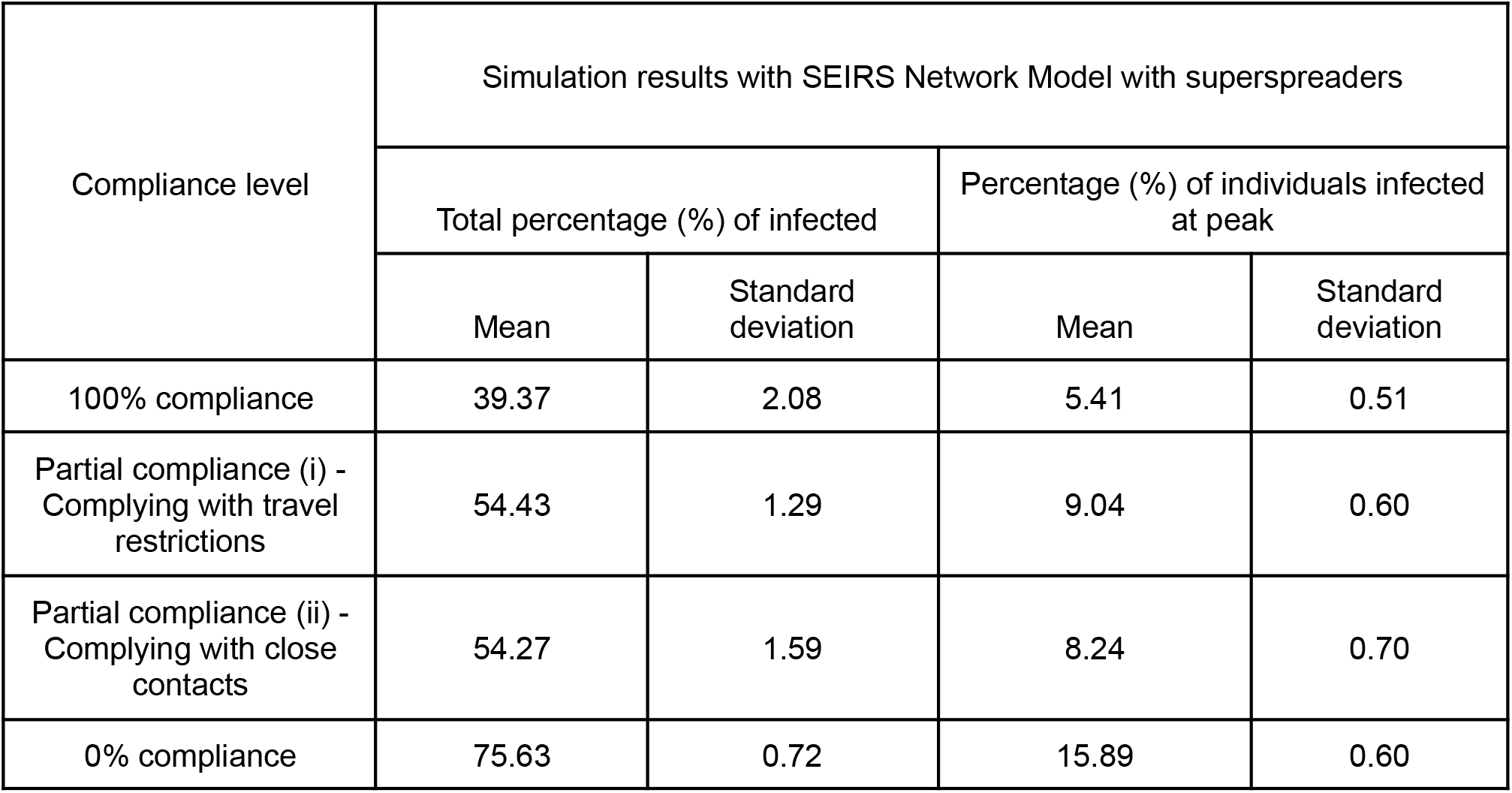
Mean and standard deviation of simulation results for total percentage of infected individuals and percentage of individuals infected at peak of heterogeneous populations (with superspreaders).

| Compliance level | Simulation results with SEIRS Network Model with superspreaders |  |  |  |
| --- | --- | --- | --- | --- |
|  | Total percentage (%) of infected |  | Percentage (%) of individuals infected at peak |  |
|  | Mean | Standard deviation | Mean | Standard deviation |
| 100% compliance | 39.37 | 2.08 | 5.41 | 0.51 |
| Partial compliance (i) -<br>Complying with travel restrictions | 54.43 | 1.29 | 9.04 | 0.60 |
| Partial compliance (ii) -<br>Complying with close contacts | 54.27 | 1.59 | 8.24 | 0.70 |
| 0% compliance | 75.63 | 0.72 | 15.89 | 0.60 |

### 3.3 Comparison of homogeneous and heterogeneous populations compliance levels

We now compare the differences in compliance levels of homogeneous and heterogeneous populations. Whilst full compliance aligns with the obvious observation of a significantly lower infection rate for homogeneous than heterogeneous populations, a striking phenomenon is seen for other compliance levels with higher total infections. Erdos-Renyi network suggests a higher percentage of infections than scale-free networks. This is consistent with results in previous literature, which show an overestimation of infections when compared to networks such as scale-free, which may be considered more realistic. It may be not intuitive at first since scale-free represent networks with nodes of high connectivity (e.g. superspreaders) [12]. It is noteworthy in any case that the infection progresses faster and a higher peak is achieved for scale-free networks, even when the total number of infected is lower. Note, however, that all networks in this work, are equally connected on average, meaning that independently of a homogeneous or heterogeneous population, the same amount of social interactions per time step will occur.

### 3.4 Effects of individual partial compliances

To further investigate the key role of superspreaders in infection spread, we now divide individuals where the top 20% of most connected individuals are considered superspreaders, as previous research has indicated that could give rise to 80% of infections [13]. We then compare the difference in effects of individual partial compliances. Parameters of partial compliances are adopted from Table 1.

We hypothesise that when superspreaders comply with travel restrictions, denoted by partial compliance (i), the results would reflect a closer resemblance to full compliance. This hypothesis stems from the fact that we believe superspreaders may be the ones who least comply with restrictions, so we are interested in studying what happens in scenarios where they comply differently to the rest of the population. Figure 10 below shows conclusive results, indicating the reduction in infections and rate by partial compliance (i).

**Figure 7.**
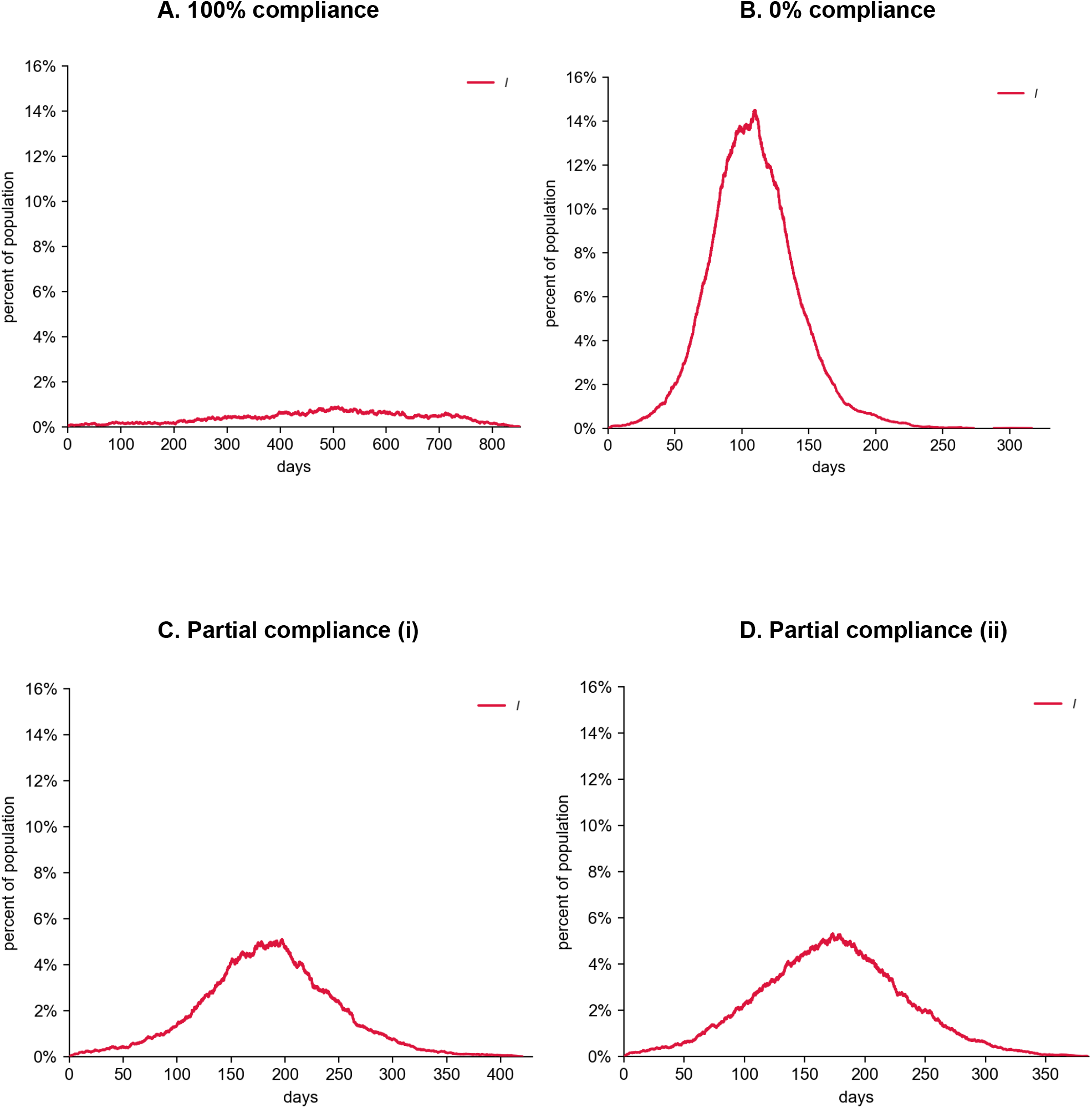
Model estimates of infected individuals per time step with different levels of compliance of homogeneous populations. Red lines represent the progression of infections. (A) Infections with full compliance. (B) Infections with no compliance. (C) Infections with partial compliance (i). (D) Infections with partial compliance (ii).

**Figure 8.**
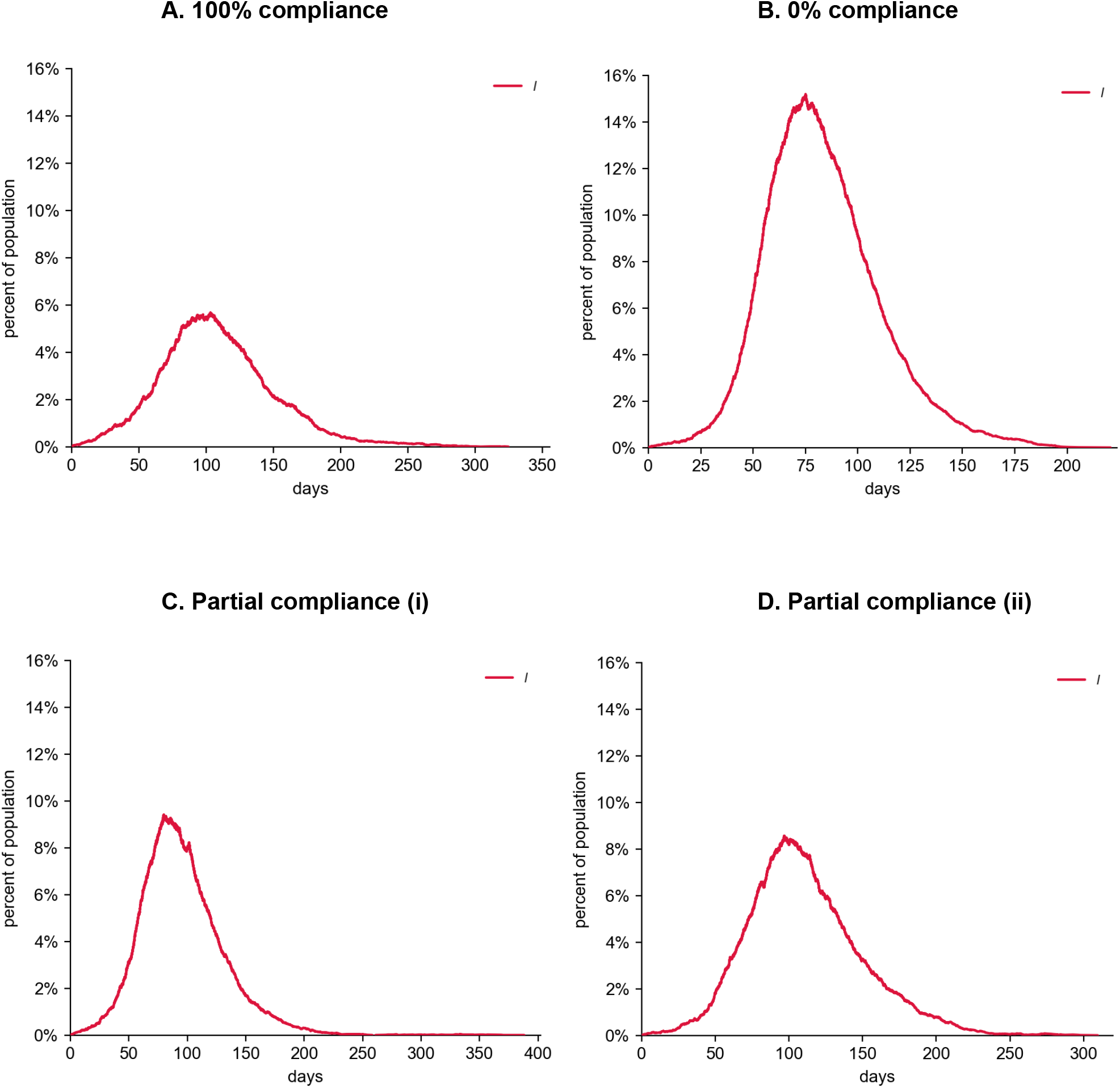
Model estimates of infected individuals per time step with different levels of compliance of heterogeneous populations. Red lines represent the progression of infections (See Data Availability). (A) Infections with full compliance. (B) Infections with no compliance. (C) Infections with partial compliance (i). (D) Infections with partial compliance (ii).

**Figure 9.**
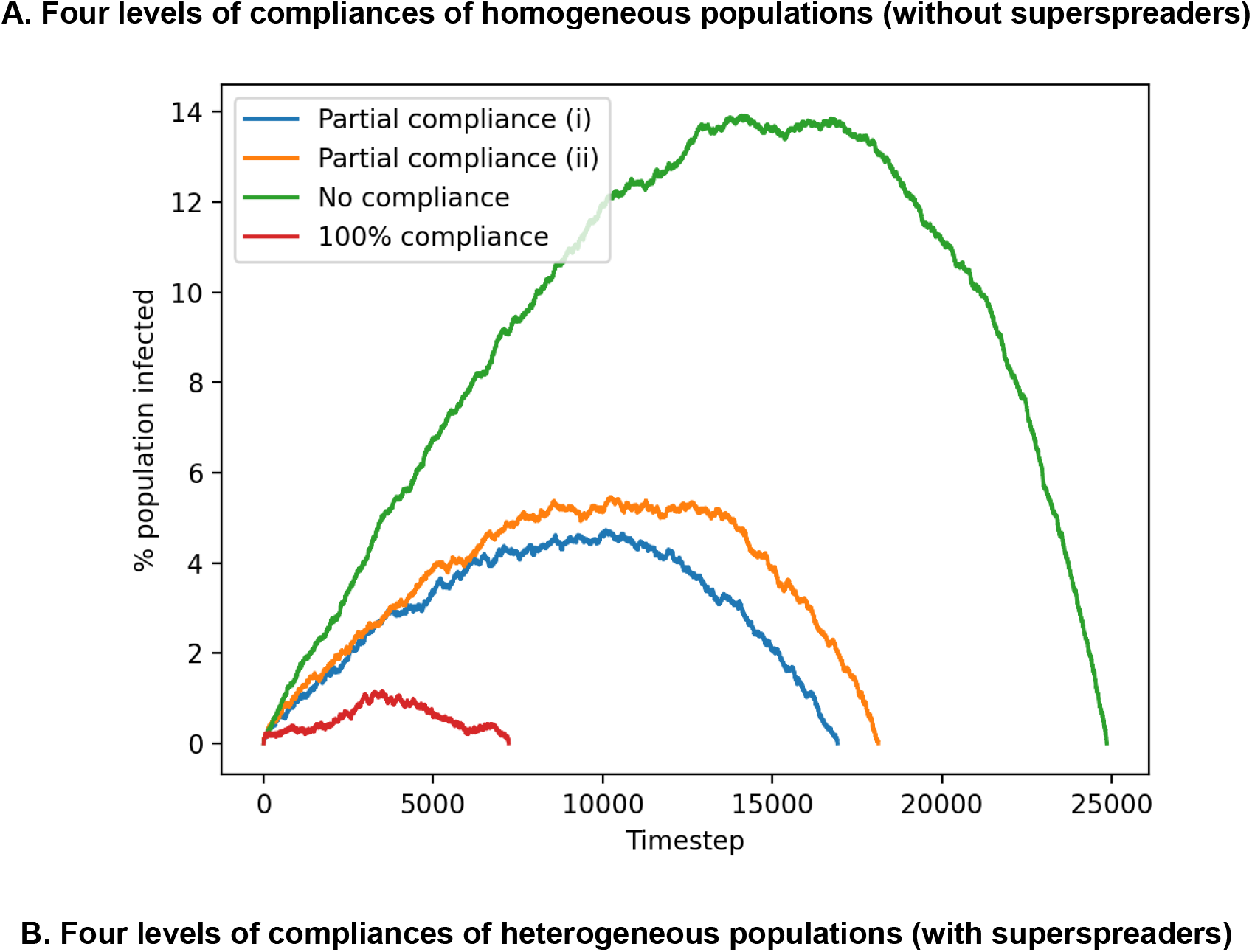
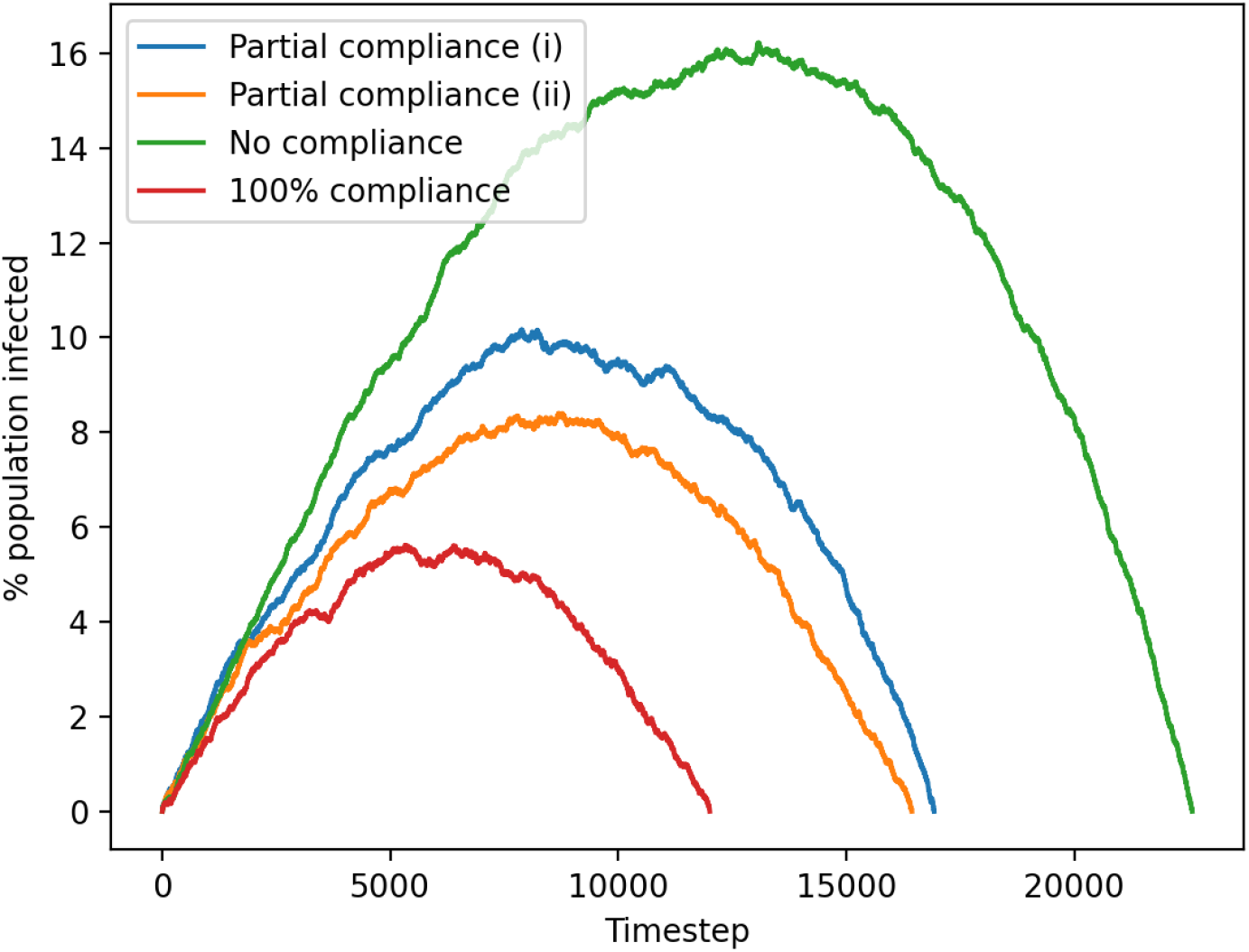
Summary of 4 compliance levels with and without superspreaders per time step. (A) Compliance levels of homogeneous populations. (B)Compliance levels of heterogeneous populations.

**Figure 10.**
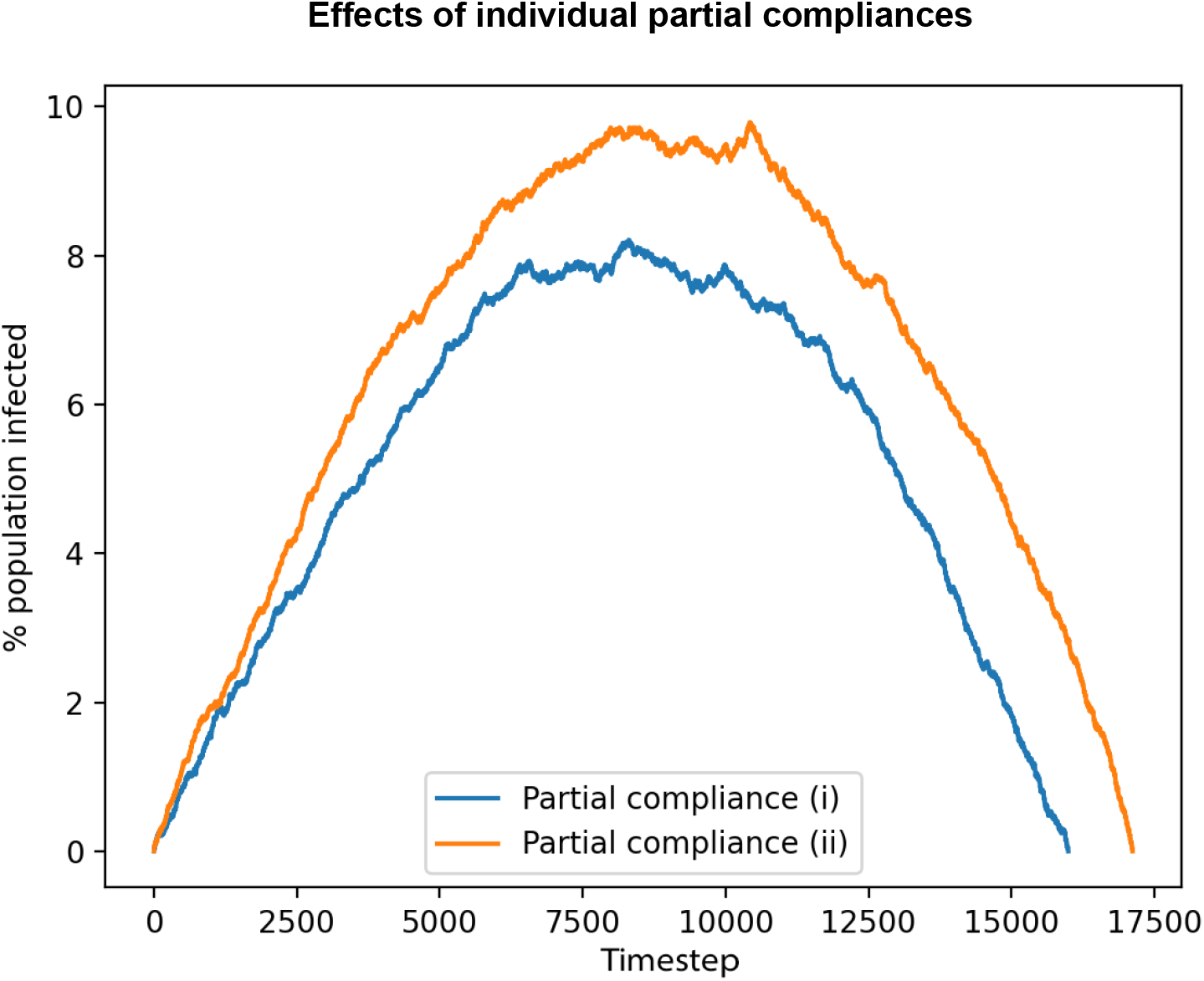
Effects of individual partial compliances per time step. The top 20% of most connected individuals are classified as superspreaders, while the rest (80%) are non-superspreaders. Blue line: Superspreaders comply with partial compliance (i) only, while the rest of the population do not. Orange line: Superspreaders comply with partial compliance (ii) only, while the rest of the population do not.

On the other hand, if superspreaders comply with having random acquaintances outdoors only (partial compliance (ii)), the number of infections increases. It is important to note that the sum of parameters p_local and p_global yield the same probability (see Table 1). This shows to a certain extent, the types of connections are critical rather than simply the amount of connections.

This is one of the examples resembling real-world scenarios, providing paramount evidence that it is crucial for superspreaders to comply with social distancing measures in order to reduce spread [14]. This allows policymakers to assess the effectiveness of currently distancing policies and evaluate for better control, such as reducing the density of crowd in indoors settings, screening of temperatures, enforcing masks and making sure places are well ventilated [15].

## 4. Discussion

Our modelling results demonstrate that compliances of homogeneous populations show a higher overall infection, except for full compliance. This shows the importance of considering more realistic social networks like scale-free in simulations aimed at policymaking [16]. Different local and global connectivities affect the spread in different manners. In particular, global interactions increase the spread and the speed of infection to the peak. However, assuming long-lasting immunity, although superspreaders make up a small proportion of the population, they could potentially slow the spread due to their high connectivity and that they are immunised quickly [17]. This is, of course, always considering the same budget of social interactions in the simulations.

An advantage of the SEIRS Network model is that it can be extended to model other diseases and predict future outbreaks with changes to parameters [18]. This allows the model to adapt to other future possibilities and scenarios, informing contact structures and providing useful references to improve current distancing measures, allowing individuals to comply in a necessary but effective manner [19]. Compared to the SEIR model, the network model highlights the heterogeneity of each node interacting with other nodes, capturing the possibility of vectors of viral transmission other than random uniform mixing [20].

As data is becoming more readily available, network models would need to be modified to collect a more detailed dataset in order to assess a range of simulated outbreaks, behaviours and policy reinforcements. Through understanding the effects of compliance of superspreaders, we can analyse patterns and target hotspots of superspreaders to implement a set of health and economic policies summarised below [8]:

- Social distancing - Complying with distancing measures leads to fewer transmission routes regardless of connection links. This can change the network structures which leads to a significant reduction in an individual’s connections. The network model used in this project can be extended to compare the difference between network level and population level by varying the parameters appropriately.
- Contact tracing, testing and isolation - Keeping track of the network of superspreaders or non-superspreaders and varying the number of contacts in the network simulation. By introducing more complex scenarios such as isolation and quarantine, a better understanding of the successful implementation of guidelines would be helpful for policymakers in future empirical work.
- Length and timing of policies - With various levels of compliance, the total and peak infections are demonstrated in the results section. It is important to adjust the duration of distancing measures based on intensity and speed of transmission. As superspreaders get infected and immunised quickly due to their high connectivity, the effectiveness of implementing intermittent policy interventions in a contiguous population will vary. If policies are lifted early, there will be continuous peaks of infection which poses consequences on economic costs. The timing of policy implementation is also worth thorough considerations due to preventative outcomes or unexpected epidemic outbreaks. A range of NPIs is advised at the beginning of SARS-Cov-2, however, multiple waves of outbreaks demonstrate the potential of continuous spread through contact networks. Thus, this model allows us to take advantage of behavioural changes and implementation of guidelines which can increase the success probability of eliminating superspreaders and reducing the basic reproductive number R_0_.
- Endogenous behavioural responses of individuals - This includes self-isolation, reducing local and global contacts. This could form one of the driven motivations of compliances due to infections or deaths observed in different contact networks. However, this model does not demonstrate the ability to predict behavioural responses of superspreaders and non-superspreaders, so future testing and development are needed to understand these patterns.

### 4.1 Limitations

Although the network model indicates a close relationship between compliance and the value of social distancing measures, an extended critique is needed in future work to understand the reasons behind various compliance levels [21]. Management strategies would need to be considered thoroughly in order to address social settings and ensure the balance between compliance and livelihood [22]. This simulation assumes long-lasting herd immunity in which further work is needed to establish detailed transmission routes providing vaccination options. The model also assumes the basic reproductive number R_0_ of 2.2 for superspreaders, which may vary as previous literature suggests it can range from 1.5-4% [23]. Therefore, further studies can be done through modelling compliances levels with varying R_0_ to explore the detailed consequences on the rate of infection spread. This also suggests infections will persist without strong effective control measures.

This model does not solely reflect the population in the UK, but rather in simulated homogeneous and heterogeneous populations. However, the main finding is to focus on illustrating the effects of compliances in a population where superspreaders exist [24]. The model reflects our understanding of the importance of compliance and adherence to government guidelines for a quicker uplift of lockdown measures [25].

Another assumption of the model framework is that we assumed a proportion of infected individuals to be symptomatic. WHO data suggests approximately 80% of infected individuals show mild symptoms [26], while other studies such as the Diamond Princess cruise reports 18% of individuals are asymptomatic [27]. Currently, there is still a high level of uncertainty in the asymptomatic population, as studies show the degree can range from 2-57% [28]. Some literature that captures the SEIR model may not have explicitly explained the differences between asymptomatic and pre-symptomatic, defining asymptomatic as the proportion of individuals not expressing symptoms at the point when they are tested positive. Further analyses would be necessary to investigate how asymptomatic individuals affect contact tracing and testing control strategies [29].

In this study, we have assumed all individuals have the same rate of transmission. However, there is no current data to show high transmission rates in adults [30]. Therefore, future studies and methods are necessary to explore that children or young adults might have the possibility of a lower transmission rate, considering the number of real-world cases stem from adults and the elderly.

### 4.2 Future work

It is crucial to quickly predict, identify hotspots and transmission patterns following the compliances of superspreaders. Further work is needed to consider more complex scenarios such as contact tracing and containment to identify high-risk and asymptomatic individuals for targeted restriction guidelines [29].

As data is more readily available, the model can be extended for analysing viral transmission by different demographics, such as compliance in different age groups or sex [23]. This could provide meaningful results for observing specific sub-groups of individuals, and identifying them using risk classification algorithms. These algorithms enable individuals to be aware of the potential rise of superspreader events and foster testing and isolation sooner. Further work in the development of contact tracing can be implemented in terms of personalised messages and reminders so individuals can avoid attending crowded events and thus reducing the number of contacts.

Following tracing, sensitive testing would be useful in identifying high transmitters with high connectivity, minimising and even eliminating hotspots of transmission routes of superspreaders [31]. Early and frequent testing would significantly reduce the infections by a fraction. These implementations would not be possible without regular environmental surveillance, which would be of great assistance in targeting superspreaders.

Besides current distancing guidelines and policies, the availability of multiple vaccines will help foster herd immunity [32]. Although the model for the simulations assumes long-lasting immunity, we can extend it to explore the possibility of vaccines in reducing peak and total infections regardless of compliances to local and global connectivity. Mass vaccination, along with a combination of enforced contact tracing, testing and isolation will allow slowing the spread and reaching herd immunity sooner. This also helps alleviate economic costs due to intermittent lockdown and closures of workplaces as a result of the pandemic.

## 5. Conclusion

The primary aim of this project is to investigate the importance of compliance of superspreaders in the successful containment of viral spread. Through simulations using the network model, we analyse the differences in speed and strength of infections between superspreaders and non-superspreaders. The main findings are that superspreaders get infected and immunised at a faster rate, and their characteristic of high connectivity produces a higher peak in scale-free networks, but a lower amount of total infections. This model at present does not show a comprehensive answer to behaviour responses and reasons for compliance, which further novel methods are needed to explore in this field of research. This study, however, provides insight for focused implementation of social distancing measures in order to target transmission routes of superspreaders, followed by effective long-term control strategies and availability of vaccines.

## Data Availability

Source code for this project can be found at data availability link.

https://github.com/debsf5/covid-compliance

## Data Availability

Source code for this project can be found at: https://github.com/debsf5/covid-compliance

## Footnotes

2 Available from: https://github.com/ryansmcgee/seirsplus/wiki/SEIRSNetworkModel-Class

